# Exploring a roadmap to achieving tobacco endgame in sub-Saharan Africa: A qualitative study among stakeholders from 12 countries

**DOI:** 10.1101/2024.09.23.24314176

**Authors:** Catherine O. Egbe, Senamile P. Ngobese, Arshima Khan, Siphesihle Gwambe, Zinhle P. Ngcobo, Stella A. Bialous

## Abstract

**Introduction:** Tobacco endgame seeks to bring an end to tobacco use or drastically reduce prevalence to <5%. Discussions about tobacco endgame and the possible strategies to achieve this goal in Sub-Saharan African (SSA) are sparse. This study aimed to explore SSA tobacco control stakeholders’ perspectives about tobacco endgame and ascertain what strategies they perceive to be suitable for the region to achieve this.

**Methods:** This was a qualitative study involving a purposive sample of 29 stakeholders interviewed via online platforms guided by a semi-structured interview schedule. Stakeholders were from academia, civil society, and government departments in 12 SSA countries. Interviews were conducted in English or French, transcribed verbatim (those in French were translated to English) and thematically analyzed with the aid of NVIVO v12.

**Results:** There is support for the adoption of tobacco endgame in SSA by tobacco control stakeholders in in the region due to the impact tobacco has on health, the environment and economy. Proposed endgame approaches for SSA were recommended to be Afrocentric which must be sensitive to cultural and regional dynamics. Stakeholders believed that the success of endgame strategies depends on political will, multi-sectoral collaboration, availability of resources, buy-in from the public and tobacco industry monitoring. Suggested endgame strategies were categorized into 5 themes: Product-focused (less addictive tobacco and regulation of novel products); User-focused (smoke-free generation, cessations supports and use of sports); Market/supply-focused (licensing of sellers, increase in taxes, control of illicit trade and alternative income for farmers); Institutional structure-focused (tobacco industry monitoring and regulation) and legislation implementation-focused (effective implementation of international treaties aimed at lowering tobacco use prevalence).

**Conclusion:** There is support for a tobacco endgame in SSA. Collaboration from various departments/ministries, and support from government and the public would be needed to make tobacco endgame a reality in SSA.

**Key Highlights:**

- Discussion on how to end tobacco use (endgame) in Sub-Saharan African (SSA) is sparse.
- Tobacco control stakeholders in SSA are supportive of tobacco endgame for the region.
- Afrocentric endgame strategies sensitive to culture and countries’ peculiarities are needed.
- User, product and supply focused, as well as institutional and legislation focused strategies have been proposed.
- Proposed endgame strategies must include alternative income for farmers and cessation support.
- Support from SSA governments and the public, and tobacco industry monitoring are needed for successful implementation of tobacco endgame in SSA.

## INTRODUCTION

Tobacco use, remains the leading cause of preventable premature death and illnesses globally.^1^ Smokers lose around 10 years in life expectancy,^2^ with an estimated one in two smokers succumbing to tobacco-related illnesses.^3^ It is estimated that tobacco will claim over one billion lives in the 21^st^ century.^4^ Therefore, tobacco endgame strategies are being proposed which are expected to drastically reduce or eliminate tobacco use^5^ to end the tobacco epidemic. The concept of tobacco endgame involves moving beyond tobacco control towards a tobacco-free future wherein the use of commercial tobacco products would either be phased out completely or their availability significantly restricted.^6^ Therefore, the tobacco endgame concept is defined as initiatives designed to change or permanently eliminate the structural, political and social dynamics that sustain the tobacco epidemic so as to achieve an endpoint for the epidemic.^7^

Definitions of tobacco endgame suggest that a key strategy in combatting the global mortality and mobility caused by non-communicable diseases is through the creation of a world essentially free from tobacco, where less than 5% of people use tobacco.^3,4,8^ Endgame strategies vary in method and aspirations, but share two underlying beliefs: (1) that the status quo is unacceptable and (2) that reducing tobacco use substantially will require something new, bold and fundamentally different from existing approaches.^1^ Proposed tobacco endgame strategies include regulating tobacco marketing, prices and profits, regulating the product to remove or reduce addictive substances and toxins, prohibition of tobacco purchasing by those born on or after a certain date, a smoker licensing system,^9^ and the elimination of the sale of tobacco products.^10^ Some authors have also proposed a complete ban on cigarettes as a tobacco endgame strategy.^11^ The targets of tobacco endgame are expected to be specific, and have measurable outcomes that would indicate an end to the tobacco epidemic in a defined geographic area (i.e. country).^12^

Countries such as Finland and New Zealand are developing plans to completely ban the sale and use of cigarettes, thus aiming to create a smoke-free society by 2050 and 2040.^13^ New Zealand introduced a ‘smokefree generation’ policy, meaning that those who are born on or after the first of January 2009,^14^ will never be legally allowed to buy tobacco products in their lifetime.^15^ Other countries which have policies indicating an aim to drastically reduce or end tobacco use include the United Kingdom, United States, Canada, Bangladesh, Scotland and Sweden.^12^ However, prior to the emergence of COVID-19, only one country, the Kingdom of Bhutan previously instituted a complete ban on tobacco.^11^ In 2004, the Kingdom of Bhutan officially instituted a national tobacco ban which specifically prohibited all sale of tobacco products and banned the use of tobacco products in public areas.^16^ However, during the peak of the COVID-19 pandemic, Bhutan reversed this ban amid concerns for cross-border smuggling and increased transmission of COVID-19. The ban on production and manufacturing of tobacco is still in place as well as a 100% sales tax on tobacco-related products.^17^ As part of measures to reduce exposure to COVID-19 or reduce the severity of COVID-19 on those who contract the disease, a few countries around the world temporarily banned the sale of tobacco products or restricted their use in public places. Notable among these countries were South Africa, India, and Botswana.^18,19^ While some researchers have attempted to draw lessons from the outcome of this ban in South Africa for endgame planning,^20^ it should be noted that the temporal tobacco sales ban was neither set to achieve endgame goals^18^ nor fits well into the definition of an endgame measure due of the following reasons. The measure: (1) was neither meant to be made permanent nor aimed at completely eradicating tobacco use (at least as indicated in South African government’s documents.^21^ 2) was not proposed as a testing ground for a future endgame strategy in South Africa. 3) did not ban the use of tobacco or nicotine products. 4) was not comprehensive to ensure other sources of tobacco and nicotine products into the country were controlled or blocked hence there were reports of illicit trade during this period. 5) was not systematically introduced as would be expected if it were to be permanent. 6) was a contingency measure to serve as one of the ways to deal with a novel pandemic which the world had little or no knowledge about as at the time it occurred.^22^

According to the World Health Organization (WHO) more than 80% of the world’s tobacco users live in low-and middle-income countries (LMICs).^3^ The Sub-Saharan African (SSA) population is particularly vulnerable as it is still in the early stages of a tobacco epidemic and has both the youngest and fastest growing population globally.^23^ The tobacco industry has also increased its marketing and production in the region^24,25^ targeting its youthful population. As a result, SSA region is projected to experience one of the highest growth in tobacco consumption in the world,^25^ with an estimated smoking rate of 37% by 2025 if current trends are not reversed.^26^ This study therefore aimed to explore what do tobacco control stakeholders perceive as tobacco endgame as well as what roadmap and what strategies do they consider to be suitable for the adoption and implementation of tobacco endgame in SSA.

## METHODS

### Design and study population

This study employed a qualitative research design using key informant interviews. The study was conducted among stakeholders of tobacco control working and residing in SSA. SSA comprise 46 countries with 44 being signatories to the WHO Framework Convention on Tobacco Control (FCTC).

### Sample and sampling technique

Purposive and snowball sampling techniques were utilized to recruit participants (tobacco control stakeholders) for the study. A total of 77 participants were contacted from 31 countries (Table S1) and 34 interviews were conducted. Data from 29 interviews with participants from 12 SSA countries were included in this study while 5 interviews were excluded because the participants mentioned that they could not speak about regional issues with regards to the topic. The inclusion criteria were adults aged 18 years and older, resident in one of the 46 SSA countries, have at least 12 months working experience in the tobacco control field, and who fell under at least one of the following categories: tobacco control advocate, tobacco control researcher, and government officials working in tobacco control.

### Data collection

Participants were first contacted via email, then telephonically and or via WhatsApp, using initial lists of civil society organizations’ contact persons provided to the researchers by the Africa Tobacco Control Alliance (ATCA) and the Africa Center for Tobacco Industry Monitoring and Policy Research (ATIM). ATCA is the regional network of civil society organizations (CSOs) and non-governmental organizations (NGOs) working in the tobacco control field in the WHO AFRO region with members in 39 countries.^27^ ATIM is a WHO Framework Convention on Tobacco Control (FCTC) observatory which conducts industry monitoring and research across the African continent. ATIM also works with tobacco control researchers, advocates and governments across the continent.^28^

Interviews were conducted via the online platform most suitable to the participant including WhatsApp, Zoom, Teams and telephone. Interviews were semi-structured and guided by an interview schedule which was developed by the researchers and derived from the key research questions guiding the study (Supplementary material S2). Participants were interviewed in either English or French languages and interviews were audio recorded. The distribution of the participants by country, sex and occupation is presented in Table 1.

**Table 1:** Socio-demographic characteristics of participants.

|  | **Category** | **n (N=29)** | **Percentage (%)** |
| --- | --- | --- | --- |
| **Country (code)** | South Africa (**SA**) | 5 | 17.2 |
|  | Nigeria (**NG**) | 5 | 17.2 |
|  | Kenya (**KY**) | 4 | 13.8 |
|  | Ghana (**GH**) | 3 | 10.3 |
|  | Benin (**BN**) | 2 | 6.9 |
|  | Senegal (**SG**) | 2 | 6.9 |
|  | Burkina Faso (**BF**) | 2 | 6.9 |
|  | Ethiopia (**ET**) | 2 | 6.9 |
|  | Togo (**TG**) | 1 | 3.4 |
|  | Zimbabwe (**ZM**) | 1 | 3.4 |
|  | Uganda (**UG**) | 1 | 3.4 |
|  | Cote D'Ivoire (**CD**) | 1 | 3.4 |
| **Sex** | Male | 22 | 75.9 |
|  | Female | 7 | 24.1 |
| **Occupation** | Non-governmental organization | 21 | 72.4 |
|  | Governmental department | 4 | 13.8 |
|  | Research/Academia | 4 | 13.8 |

### Data analysis

Recorded interviews were transcribed verbatim, and the French transcripts were translated to English. Thematic analysis was conducted on the qualitative data with the aid of the software NVivo version 12. The analysis followed the Braun and Clarke’s six-step framework for conducting thematic analysis (namely familiarizing yourself with the data; generating initial codes; searching for themes; reviewing themes; defining and naming themes; and producing the report).^29^ Suggested endgame strategies were categorized guided by the ‘policies with potentials to achieve endgame’ published by Puljević and colleagues (product focused, user focused, and market/supply focused, institutional focused). COE, AK, ZN and SG initially coded a sample of transcripts, after which, AK, ZN and SG completed the analysis. Completed analysis was reviewed by COE and SPN afterwards. SAA helped with resolving disagreements with interpretation and categorization of themes during this process.

### Ethical considerations

Ethical clearance to conduct this study was obtained in April 2021 from the South African Medical Research Council Human Research Ethics Committee (Protocol ID: EC014-4/2021). Information sheet/consent forms (ICF) were used to communicate details about the study in English and French. Each participant was requested to read through the ICF, sign and return same to the project team as confirmation of their intention to voluntarily participate in the study. Participants were informed about their right to discontinue or withdraw from the study at any time and were assured of anonymity and confidentiality throughout the study. Participants were also informed that all information shared or published would be de-identified except for country names. Code names have been used to ensure anonymity. Audio recordings and transcribed data have been stored in password-protected laptops only accessible to the principal investigator and co-authors.

## RESULTS

Findings were grouped into five categories with their associated themes and subthemes. These categories included perception about the concept ‘tobacco endgame’ and its importance for SSA; perspectives on how SSA should approach tobacco endgame; roadmap to the implementation of tobacco endgame in SSA; facilitators to the adoption and implementation of tobacco endgame in SSA and suggested tobacco endgame strategies for SSA. There was a general perception that effective implementation of the WHO FCTC in controlling tobacco is an essential roadmap towards tobacco endgame in SSA because the strengthening of tobacco control measures would adequately prepare SSA countries for the adoption of endgame strategies. A summary of the results is presented in Tables 2 and 3.

**Table 2:**
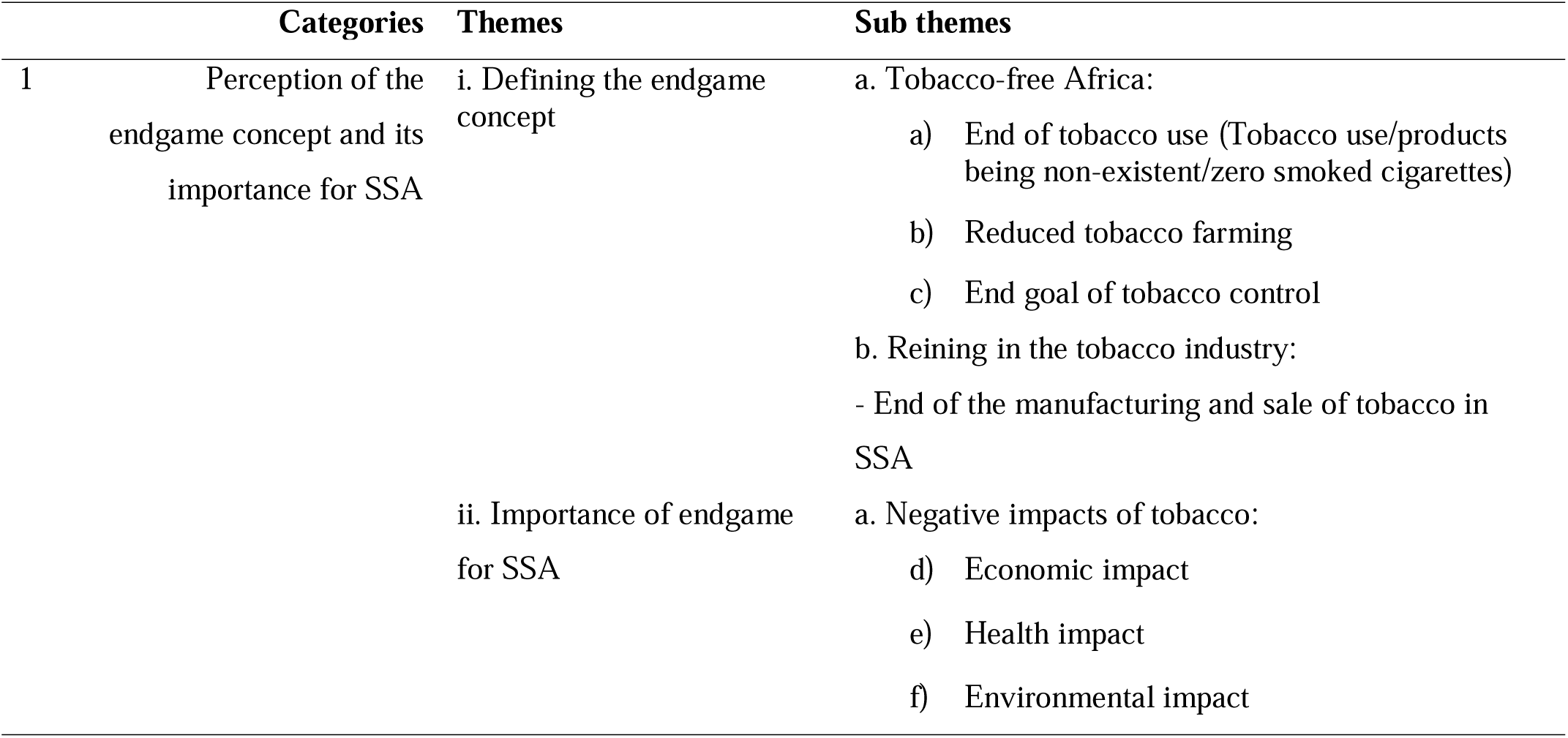
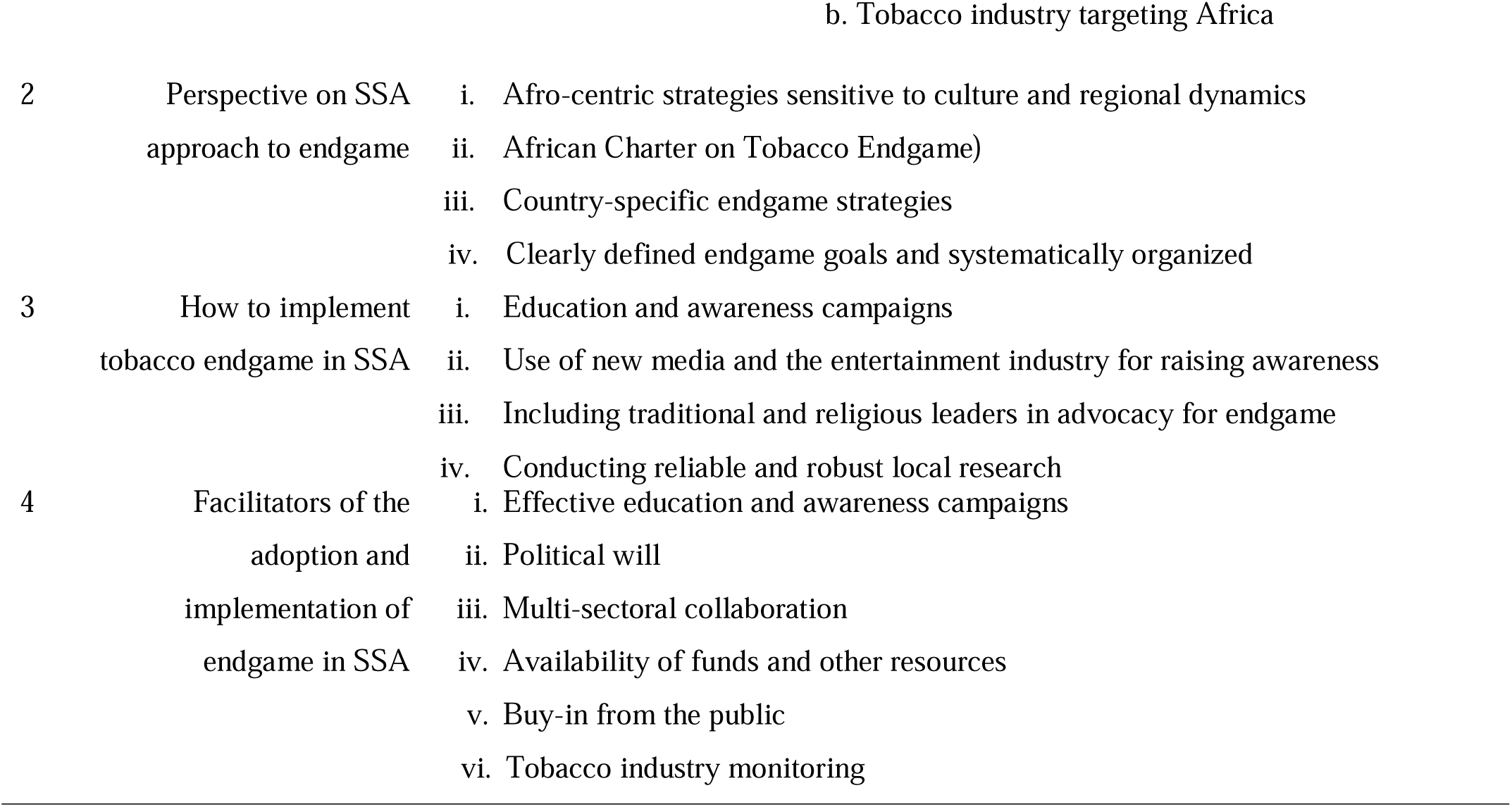
Themes on stakeholders’ perceptions on the concept, approach and facilitators of tobacco endgame in Sub-Saharan Africa.

**Table 3:**
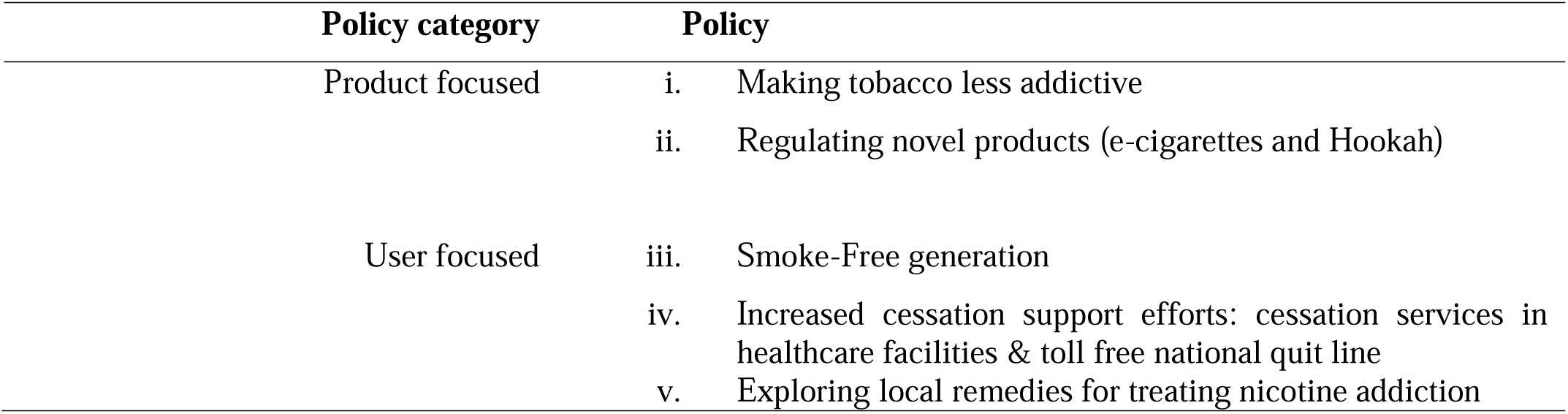
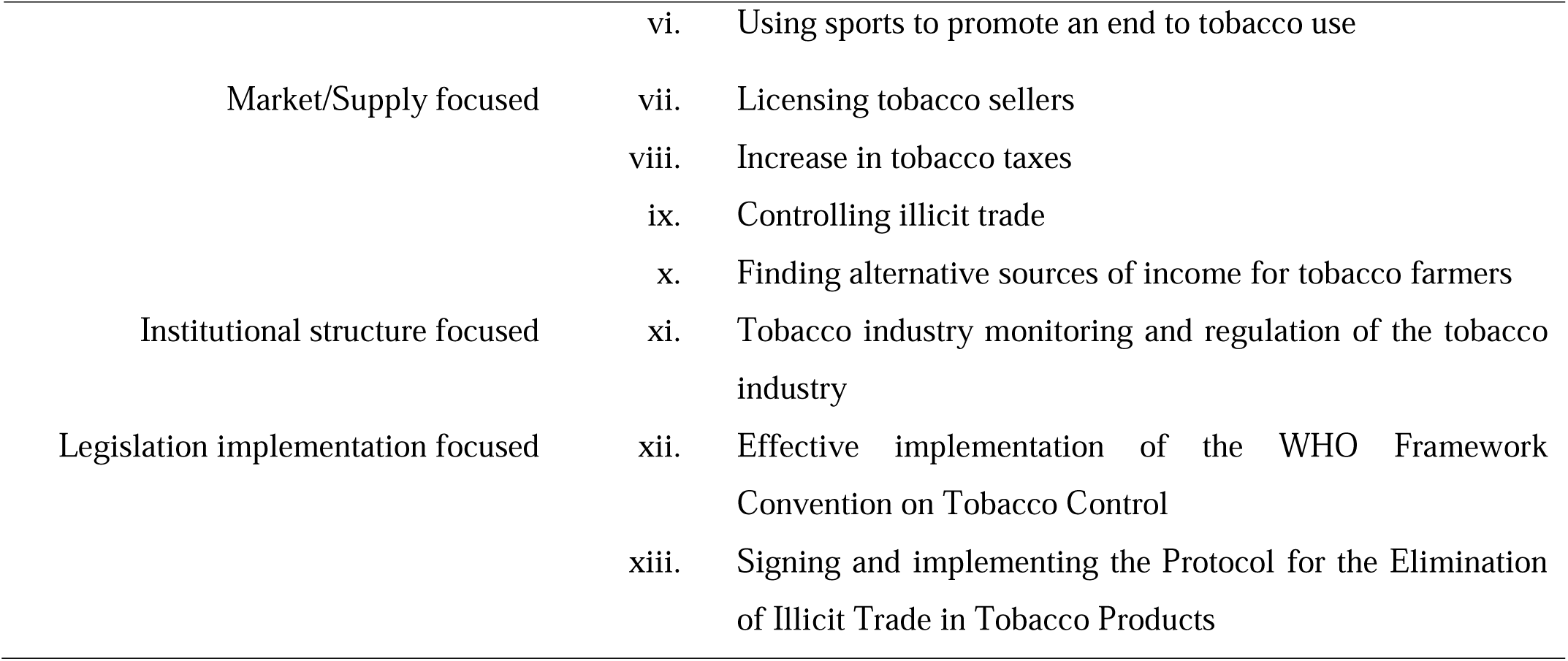
Policies with potential to achieve tobacco endgame in SSA.

| **Policy category** | **Policy** |
| --- | --- |
| Product focused | i. Making tobacco less addictive |
|  | ii. Regulating novel products (e-cigarettes and Hookah) |
| User focused | iii. Smoke-Free generation |
|  | iv. Increased cessation support efforts: cessation services in healthcare facilities & toll free national quit line |
|  | v. Exploring local remedies for treating nicotine addiction |

|  |  |  |
| --- | --- | --- |
|  | vi. | Using sports to promote an end to tobacco use |
| Market/Supply focused | vii. | Licensing tobacco sellers |
|  | viii. | Increase in tobacco taxes |
|  | ix. | Controlling illicit trade |
|  | x. | Finding alternative sources of income for tobacco farmers |
| Institutional structure focused | xi. | Tobacco industry monitoring and regulation of the tobacco industry |
| Legislation implementation focused | xii. | Effective implementation of the WHO Framework Convention on Tobacco Control |
|  | xiii. | Signing and implementing the Protocol for the Elimination of Illicit Trade in Tobacco Products |

### Perceptions about the concept of tobacco endgame and its importance for SSA

#### Definition of the “tobacco endgame” concept

The participants were asked to explain what they understood by the concept ‘tobacco endgame’. Participants definitions were grouped into two main themes: tobacco-free Africa (end of tobacco use) and ending the tobacco business. Findings show that some participants already have a working vision of the region and their countries being tobacco-free in the future. Slogans mentioned by a participant to express this vision include “tobacco-free Africa” and “tobacco-free Kenya”.

> *“We talk about the tobacco free Africa, in Kenya, we say ‘tobacco free Kenya’. Meaning that we are looking forward to a situation whereby we will not be having tobacco at all. And that is something…the vision is very valid.” (KY_01)*

For other participants tobacco endgame meant ***the end of tobacco use*** in all its forms and is the end goal of tobacco control.

> *“Tobacco endgame would assume that we will arrive at a point whereby the consumption of tobacco in its different forms that we know will no longer exist, and that more and more the population especially young people will turn away from tobacco in all its forms, when I talk about all its forms as for example nowadays, in terms of cigars, cigarettes, cigarillos, tobacco in the forms of new products such as shisha and others, in all its forms it will end”. (CD_01)*

However, other participants understood the concept to mean a way *to **rein in or end the tobacco industry*** which was seen as a business that propagates the tobacco problem. This implies an end to the manufacturing, sale, use and farming of tobacco.

> *“Tobacco endgame, actually you know, I have one slogan… when I am talking about the industry, this is the industry that should die. Tobacco endgame is to me to finish the tobacco business and to make the whole globe tobacco free, so that means there isn’t any tobacco issue or product in the whole earth. (ET_02)*

#### Importance of tobacco endgame for SSA

All participants agreed that tobacco is negatively impacting the SSA region. Three subthemes emerged as the reason the complete eradication of tobacco is important for the region: the negative impact of tobacco, SSA’s fragile healthcare system, and tobacco industry targeting Africa.

The negative impacts of tobacco were grouped into the economic, health and environmental impact of tobacco. The ***economic impact of tobacco use*** on health and health systems in SSA was of concern to participants. This impact includes the financial cost of treating tobacco-related illnesses which burdens the already weakened economy as well as the strain on the fragile health system. A participant stated,

> *“…it has become so imperative because we are a struggling and a developing country and therefore, we do not have the luxury of better health care, we do not have the luxury of a better condition to manage crises as compared to the western world and that is why it is very important, it is a matter of urgency that we step up the campaign and this endgame campaign, in order to prevent ourselves for the continent to get into a situation of dire health and economic situation….” (GH_02)*

A participant described the net financial loss governments suffer as a result of tobacco when compared with the revenue generated from the industry.

> *“The tobacco industry will talk about taxes that they bring to a government, but when you work it out the government spend more in treating tobacco related diseases, and also the vulnerability that is brought as a result of tobacco use.” (KY_03)*

Participants also mentioned that tobacco leads to serious ***health implications*** which are preventable, and which then puts additional burden on the already burdened health system in SSA, as seen especially during the peak of COVID-19. They noted how the region is facing multiple challenges with the ever increasing communicable and non-communicable diseases.

> *“Looking at the healthcare system, the health infrastructure, you realize that in Africa, we are still battling with TB and HIV/AIDS and Malaria, we are still battling with them. COVID has exposed us even more, we don’t have the health facilities, it’s not easy to admit as citizens, we don’t have it.” (GH_01)*

> *“We are concerned about this [tobacco] because of the impact it has on health, and on the health systems. Mind you, in Africa, most countries, their health systems leave much to be desired, you know, they are not well resourced in terms of person power, they are not effective and efficient, they don’t have the systems, they don’t have the tools to enable them to run health systems in an effective manner. And so, we cannot afford unnecessary diseases affecting the people who are really sick with the tobacco related diseases affecting the systems, health systems unnecessarily. (SA_10)*

The ***environmental impact of tobacco use*** which includes pollution of the environment was also mentioned a reason endgame strategies are important to implement in SSA.

> *“Tobacco pollutes the environment, especially in agriculture. They use several chemicals like pesticides, and they will have an impact in the future, they are cancerous, they will cause cancer in the future and such chemicals are transferable in nature, they will transfer from the soil, water, and other vegetables, then they will be eaten by the people, you know, it’s a continuum.” (ET_02)*

#### Tobacco industry targeting Africa

Another reason participants gave reasons why tobacco endgame strategies are important for SSA is the targeting of Africa by the tobacco industry, especially young people in the region. Participants believed that ending tobacco use would protect the youth within the region.

> *“[We need to end tobacco use] …because the tobacco industry is targeting the youth and they want to make Africa their major market.” (NG_01)*

### Perspectives on SSA approach to tobacco endgame

Stakeholders were asked if they believed that unique strategies were needed to end tobacco use in Africa. The features of SSA tobacco endgame strategies as suggested by stakeholders should be ‘Afrocentric’ and sensitive to culture and regional dynamics; in the form of an Africa Charter on tobacco and health; be country-specific, have clearly defines goals and be systematically organized.

Stakeholders proposed that SSA endgame strategies must be ***Afrocentric***, meaning that these strategies must cater to the African context and be sensitive to African culture and regional dynamics given the diverse nature of the region.

> *“Ethiopia has a culture that discourages this product, that means it is going to violate the culture of the community if it is expanding in the country…maybe we can use cultural means to implement the endgame.” (ET_02)*

> *“Like I said earlier on, every continent has its dynamics, Africa has its dynamics, and even in the region of Africa, the sub-regional countries have its own dynamics, southern Africa, eastern Africa, northern Africa, western Africa, central Africa, even though it all belongs to one region, there are dynamics, you get it. That is where strategies are very key and strategies that are to be implemented by the endgame to achieve this where we will end tobacco in Africa, must also be looked at based on the sub-regional dynamics and that is where strategies can be implemented effectively to achieve the results. That is why I said if Africa is able to have its own strategy it must be even disseminated to sub-regions and each nation will be able to have its own strategies out of those ones.” (GH_02)*

Furthermore, a participant proposed an ***African charter*** for tobacco and health that would encompass a regional position on tobacco.

> *“To actually have an African position on tobacco and health. We do need to have like an African Charter of Human Rights, for instance, which flowed directly from the UN Charter on Human Rights. Can we begin to work for an issue position resolution on tobacco use on the continent?” (NG_02)*

Participants posited that countries within SSA, although similar, are unique because some countries are dependent on tobacco farming. They mentioned that while a regional framework for tobacco endgame is possible, the same endgame strategies may not be applicable for all countries within the region. Such frameworks should take into consideration whether the country is a tobacco growing, manufacturing, or importing nation. ***Country specific endgame strategies*** were therefore proposed.

> *“I am not too sure I can use the word unique. However, I would want to use the word; strategies that respond to the needs, maybe that’s where unique comes in, unique to the environments of each country, I can put it that way. Because countries differ politically, countries differ in the way they are structured economically, and the framework perhaps is the same. We can have the same framework, but what could differ is how the strategy is executed, because each strategy has to respond to a particular environment. Of course, there would be similarities, but the strategy is the policy, and if we look at the policy, and you then have your regulations. If you look at regulations you need to have all those functional elements of policy execution and of course that could differ, and then of course the implementation will differ country by country.” (SA_10)*

> *“I think it’s going to be country dependent on how those countries are, how they prioritize tobacco in the first place and what they do in the area of tobacco control; are they growers, are they manufacturers or are they just importers? It’s a number of things that needs to be considered, not just one blanket solution.” (NG_03)*

Participants also suggested that strategies must have ***clearly defined goals and be systematically organized***, to ensure the effectiveness of such measures.

> *“To reduce tobacco use to the barest minimum or even stop it, it has to have a coordinated approach, it has to be a systematically organized strategy. It’s not just to try this today. So, you need a coordinated endgame strategy to be able to achieve the goal that you need. If not, we’ll just be doing all sorts and I don’t know whether it will be effective. So, for effectiveness, then we need the endgame strategy.” (NG_01)*

### Road map to the implementation of tobacco endgame in SSA

Findings from this study revealed that while embarking on tobacco endgame, some important steps to take to ensure its success include public awareness programmes; use of new media and the entertainment industry to raise awareness; including traditional and religious leaders in advocacy and conducting reliable and robust local research to inform policies.

***Public awareness*** (through public education and communication) was seen as a type of strategy that SSA can use to educate people about the dangers of tobacco, importance of health, and decrease the consumption of tobacco in the region.

> *“…there should be massive education on the continent about the dangers of tobacco use.” (NG_02)*

Participants stated that these campaigns should be targeted at adolescents and women due to the increasing tobacco use among these populations.

> *“more and more we really note that there are young girls who are now subscribed to tobacco… if we develop information and education programs and if we involve the grouping of women’s training and youth movements, I think that we can actually succeed in redressing the use of tobacco although it is very difficult to eradicate strictly the use of tobacco in our countries.” (SG_02)*

The ***use of new media*** and the entertainment industry for awareness as well as ensuring smoke-free movies were also mentioned as a strategy to discourage smoking in the population towards ending tobacco use.

> *“…use the entertainment industry to set a template for, to set a standard, which you get to stop smoking in movies, and for celebrities to now also serve as role models to discourage smoking…” (NG_04)*

Additionally, stakeholders suggested that there is need to ***include religious and traditional leaders*** in the advocacy to end tobacco use given the high regard Africans have for these two institutions.

> *“Africans are people who actually uphold their culture in a high esteem, and I believe that if we are able to get our messages across various religious leaders and traditional authorities, where we are able to make them understand and for them to also buy into our campaign, when the messages and the advocacy and education is coming from the religious leaders, is coming from the traditional authorities, where people hold in high esteem. I believe that it is one strategy that can help a lot in order for us to be able to end this tobacco consumption.” (GH_02)*

Participants also emphasized the need for more ***reliable and robust local research*** within the SSA region to inform endgame policies. According to one participant, there is need to develop and rely upon data generated from African countries to inform solutions to regional problems.

> *“Africa, we need our own material…we need to start telling our own stories, not using the stories of beyond borders, to try solving a very unique matter on our own soil, on our land…” (NG_02)*

### Facilitators of the adoption and implementation of tobacco endgame in SSA

Stakeholders proposed some facilitators to achieving successful tobacco endgame adoption and implementation in SSA. These factors include effective education and awareness, political will, multi-sectoral collaboration, availability of funds and other resources, buy-in from the public, and tobacco industry monitoring. Participants mentioned that ***education and awareness*** must be hinged on reliable and unbiased data and media campaigns which are more likely to catch the attention of policy makers.

> *“So, I believe that… carrying out studies and working with the national statistical agency of the scientific and demographic of the country which regularly conducts research…will be possible for us to tell the authorities that govern us for example what is needed for each area…” (SG_02).*

***Political will*** from regional, subregional bodies and national governments as well as decision-makers was also identified by participants as one of the main factors that would fast-tract tobacco endgame in SSA.

> *“Something that also came to me is government commitment or political will. I really think that is the most important factor, because once these things are in place, for example AU [African Union] should pick this up as top priority and cascade it down to other countries so they can also take this as a priority, there will be an endgame in a short while because the commitment will be there.” (NG_03)*

Another factor mentioned was ***multi-sectorial collaboration***. Participants suggested that a unified approach among all stakeholders and government departments will help end tobacco in SSA.

> *“No human work is perfect, but I think that the will counts, if the will is there at the regional and national level even the involvement of all actors such as civil society and others, if everyone gets on with it, nothing is impossible, we often say that together we go far, so if we get together, it is obvious that we will go far.” (CD_01)*

Stakeholders emphasized the importance of adequate ***human and financial resources, buy-in from the public*** and ***the media*** to determine the success of tobacco endgame in SSA.

> *“The mobilization of resources…I am talking about human resources, it is necessary that there are more actors who are interested in the theme, we also need financial support to always innovate in terms of actions.” (CD_01)*

> *“…media buy in. if there’s media interest, if the media is ready to talk and write about it, that is a good factor…” (GH_01)*

Lastly, ***tobacco industry monitoring*** was mentioned as an important factor that will also determine the success of tobacco endgame in the SSA region. According to some participants, most challenges against the endgame will likely stem from tobacco industry interference.

> *“We need to continuously monitor the tobacco industry…So, the tobacco industry is working 24/7, so we also need to be watching them 24/7 and doing something about it, and building operations including the media that will continue to embarrass the industry, to put the information out there to contradict the lies that they are putting out there.” (UG_0)*

> *“And of course, that goes without saying that tobacco interference should be stopped or reduced to the barest minimum. These factors will go a long way in helping tobacco control and finally ensuring that we prevent younger people, non-smokers from taking on tobacco smoking.” (NG_01)*

### Policies with potential to achieve a tobacco endgame in SSA

We used the four groups of ‘policies with potential to achieve a tobacco endgame’ published by Puljević and colleagues which include product focused, user focused, market/supply focused and institutional structure focused,^12^ to categorize stakeholders’ suggested endgame strategies for SSA. An additional category was added to this group of policies which covered actions geared towards reducing tobacco use prevalence. Not all groups of policies under each category were identified in this study, however, additional policies suggested by stakeholders were added to the various identified policy categories (Table 3).

#### Product-focused strategies

Endgame strategies focused on tobacco and related products included a reduction in the addictiveness of tobacco products and the regulation of novel products like e-cigarettes and re-emerging tobacco products like Hookah. Participants saw these as policies with the potential to end tobacco use.

> *So, I think for me one strategy that we can use, that I really liked when I heard about, is making tobacco less addictive. Reducing nicotine content and move it to less addictive rates. because for me stopping it completely will be difficult, so progressively cutting down on the addictiveness of tobacco over time, that is what Africa should be doing. (SA_09)*

> *“Regulating e-cigarettes and all kinds of novel tobacco products and that’s going to have a big impact on the e-cigarette market.” (SA_08)*

#### User-focused strategies

In addition to the regulation of products-related strategies, stakeholders mentioned the need for a ***smoke-free generation*** policy. This is a strategy entailing putting in place a legislation where children born in, or after, a particular year can be prevented, by law, from buying tobacco products.

> *“Part of one of the endgame strategies is the so called ‘smoke free’ generation. I think in the Netherlands, they had the strategy, if that’s still the case, everyone born in the 25th century not a single one we want to see smoke, so what that means is that in 50, 60, 70 years’ time; the smoking generation would have died out.” (SA_08)*

The importance of having ***cessation services*** in healthcare facilities and a national quit line was noted as a strategy to end tobacco use in SSA.

> *“The other strategy could be cessation. One; once they have the addiction…, they have a right to have this intervention to be free from this addiction. So, the service should be integrated within the health services for instance in the primary health care system.” (ET_02)*

> *“I think we can do more in the African region as a whole, the cessation and the addiction part is something we need to prioritize over time, but we are already seeing other countries start, and those services will be available.” (NG_03)*

Another user-focused strategy proposed was ***exploring local remedies for treating addiction*.**

> *“And also, there should be local based measures especially as I have mentioned, using specialized local available material to heal that [nicotine] addiction, So, overall implementation of cessation services which are local to the country or continent is very, very important.” (ET_02)*

Lastly, stakeholders were of the view that when young people are encouraged to take up ***sports and*** join ***fitness clubs*** this could help to dissuade them from using tobacco.

> *“I believe that sport can be a means for us to fight against tobacco…. I believe that if we ensure that at an early age, citizens are introduced to sport and citizens have the opportunity to subscribe to sports even if there are not enough of them at the moment and if there are opportunities that are put in place, I believe that this will be important. In Senegal we notice more and more fitness clubs where there are people especially young people and adults who now subscribe to sport. Personally, I believe that in this area of the development of sports practice we can also contribute effectively because we remain positive in the fight against tobacco.” (SG_02)*

#### Market/supply-focused strategies

One participant mentioned that tobacco products like Hookah is available everywhere and there is need to have control over this by licensing sellers.

> *“Think about hookah and the hubbly bubbly, it’s to freely available. There are no licenses, there is no control over it, and also it is an addiction concern.” (SA_08)*

Some participants believed that if ***tobacco taxes are increased*** significantly, this could help to reduce tobacco use drastically.

> *“So, I believe that if we continue to increase the taxes, I mean who knows we can reduce tobacco use to decrease to maybe single digits, below 10%” (SA_04)*

The ***control of illicit trade*** was described as an important strategy to reduce the availability of tobacco products. For example, a participant explained that the track and trace system was introduced to fight against illicit trade in Kenya and has proven to be effective.

> *“The Kenya Revenue Authority, fighting illicit trade, introduced a track and trace program. That is a win. It is to ensure that every tobacco that is sold in the country is a legal product. It has not been snuck through the borders; it has not been taxed. So, it is generally a way or a mechanism that has been created by the Kenya Revenue Authority that ensures that we, that all the tobacco that is sold in the country, has been taxed, is here legally and that is important.” (KY_03)*

Some participants suggested that any endgame strategy planned for SSA must include as a key component addressing how to help ***tobacco farmers transition into farming other crops*** to protect their means of livelihood.

> *“…there won’t be any endgame in Africa without the policy to move tobacco farmers, from cultivating tobacco to other wholesome products of their livelihoods that are already dependent on tobacco farming in Africa. So that also should be a key component of our endgame strategy.” (NG_02)*

> *“We in order to implement this endgame policy and to bring it about successfully, need to encourage and in fact, get people to support and sponsor farmers to change from tobacco into more profitable crops, there are many” (SA_06)*

#### Institutional structure-focused strategies

Institutional structure focused strategies mentioned by the participants include ***tobacco industry monitoring and regulation of the tobacco industry***. Some participants noted that the tobacco industry continues to manipulate laws and exploit SSA for profit, hence it is important to continuously monitor this industry to address tobacco use in the region.

> *“And so, the industry must be regulated, the industry must be monitored, the industry must be forced to report, even including their revenues, if their revenues keep escalating, it means something is wrong around tobacco control interventions, it means they are still getting more out of a product killing people, you see. So, the industry is a major threat and what we need is strong and robust regulation of the industry.” (SA_10)*

> *“If we can continue to monitor what the industry is doing, we will be able to respond. I think that will make a difference for Africa.” (UG_01)*

#### Legislation implementation-focused strategies

For some participants, endgame strategies were important but not a matter of urgency for now because according to one participant there is need to ***lower the prevalence of tobacco use*** in the region before an endgame strategy is considered. To lower the prevalence of tobacco use was considered as a pre-requisite for countries in SSA to embark on an endgame path and stakeholders also proposed that this can be achieved by effective implementation of international treaties like the WHO FCTC and the Protocol for the Elimination of Illicit Trade in Tobacco Products (ITP). Some participants also believed that these treaties in themselves if effectively implemented could end tobacco use in the SSA region.

> *“You only start thinking of the endgame strategy if your smoking prevalence has decreased a lot, and is becoming very low, and starting so low that you start thinking about the 5%. If your smoking prevalence is still 15 to 18%, as is in South Africa’s case, I don’t think it’s the right time to be thinking about endgame strategies as yet.” (SA_08)*

> *“We fully implement all the provisions of the FCTC, the provision of our tobacco control laws. So, far this is how I see the end of tobacco, the endgame in Africa.” (SG_01)*

One stakeholder from South Africa particularly mentioned the need for the ***signing and implementation of the Protocol to Eliminate Illicit Trade in Tobacco Products*** ^30^ by a country like South Africa where illicit trade is common as a way to reduce smoking prevalence.

> *I think important in South Africa is the illicit trade…. I think one of the key things to do on[in] South Africa, is to try and eliminate illicit trade, but not only just signing it because I think sometimes countries, or people in general, will just get into the habit of just signing things and putting them on paper, but not implementing them, so it is not only the matter of signing the protocol, but actually try and implement so things like the illicit trade and tax, make sure that we implement.(SA_09)*

## DISCUSSION

To the best of the knowledge of the authors, no study has been conducted to explore the perception of tobacco control stakeholders about the concept of tobacco endgame in Africa to date, making this study the first to do so. The results presented how stakeholders in the SSA region perceived tobacco endgame. Their understanding of endgame, though varied, were in line with previous definitions of moving away from controlling tobacco use towards ending it completely and envisioning a tobacco-free future^31^ for their country and the region.

While there are discussions and policies already being put in place to move tobacco control policies and interventions towards an endgame plan in some parts of Europe and America, there is no data showing that any country in SSA has considered this vision.^12^

SSA tobacco control stakeholders do believe that tobacco endgame strategies are important because of the negative effects that tobacco has on the environment, health, and the economy. This importance is especially key given that the SSA region is projected to experience one of the highest increases in tobacco use prevalence by 2025 if no action is taken.^26^

Findings show that stakeholders were particular about the nature of the endgame that would be suitable in the SSA regions. Afrocentric approaches which should be particularly designed to fit the context and culture of Africa and regional dynamics as well as the involvement of religious and traditional leaders who are highly respected public figures in the region would be more suitable for SSA. This finding supports the assertion by^32^ that for endgame approaches “different solutions will prove possible in different places, and may unfold in unique ways.” (p. 43).

Our findings also reveal that the stakeholders believe that success of the tobacco endgame in SSA would depend on various factors, from political will to public support. Political will is instrumental in establishing legislation and successfully implementing tobacco endgame strategies.^5^ In Uruguay, key politicians supported tobacco control which led to a decrease in prevalence by 7% in 3 years.^5^ Research has shown that the public does support efforts to decrease or end the use of tobacco if properly defined and communicated by the government.^31^

As previously mentioned Puljević and colleagues described some policies that have the potential to achieve the endgame grouped in four categories; product focused, user focused, market/supply focused and institutional focused.^12^ In addition to these key policies, this study also recommended a legislation implementation focused approach. This approach, which should primarily focus on effective and full implementation of the WHO FCTC as well as signing the ITP will focus on reducing prevalence which was seen by some stakeholders as a pre-requisite for embarking on an endgame plan.

While some stakeholders in this study considered the smoking prevalence of many SSA countries too high for the consideration of embarking on an endgame approach, it should be noted that there are some countries in the region who already have a low prevalence of less than 10% and could be ready to take on an endgame approach to end tobacco use. Examples include, Ethiopia and Tanzania prevalence of 5% ^33^ and 8.7%^34^ respectively. These and many other SSA countries with low prevalence could adopt endgame strategies, provided there is wide public understanding and support across various groups, and use of evidence-based strategies.^5,35^

Important for consideration is the fact that even in countries which have embarked on endgame, tobacco control measures are being implemented concurrently with tobacco endgame measures. This implies that tobacco control and endgame measures are not mutually exclusive and should be implemented hand in hand as weak tobacco control implementation would be inimical to the effective implementation of tobacco endgame measures.

Our study has shown that despite the different elements needed by different SSA countries to possibly embark on an endgame journey, our findings suggest that there is support for tobacco endgame among stakeholders of tobacco control in SSA. However, further research is needed to explore the general public’s understanding of the endgame concept as well as its purpose to curb the tobacco epidemic in SSA.

### Limitations

This is a qualitative study which generated data from key informant interviews. While consistent with achieving our aim, it limits our ability to generalize our findings to the entire tobacco control stakeholder community in SSA. The stakeholders in the field of tobacco control, however, were carefully selected to cut across at least one-third of the countries in SSA and various sectors including academia, advocacy, and government. The selection criteria also ensured that these stakeholders have worked in the tobacco control field for at least one year prior to the study. Further research should include a higher number of stakeholders in a quantitative survey to ascertain if this support is widespread.

## CONCLUSION

The study reveals that there is support for tobacco endgame among stakeholders in SSA. Tobacco control stakeholders also believe endgame strategies are urgently needed because of the detrimental effects caused by tobacco use and industry activities targeting young people in the region. Proposed endgame measures which could facilitate the eradication or significant decrease in tobacco use in SSA were recommended to be Afrocentric, sensitive to the cultural and regional dynamics, systematic and with clearly defined goals. For the endgame strategies to succeed in SSA, collaboration among various government departments, stakeholders and support from the public was recommended. Product, user, market/supply, institutional structure as well as legislation implementation focused policies were suggested. Findings from this study can be used to continue the conversation and inspire regional consideration for tobacco endgame in the SSA region.

## Authors’ contribution

COE conceptualized the study, COE, SPN, AK, SG, and ZN analyzed the data, SPN, AK, SG, and ZN wrote the first draft. COE, SPN, AK, SG, ZN, SAB contributed to the interpretation of results and COE and SAB critically reviewed several drafts of the paper. All authors approved the final draft for publication.

## Acknowledgements

Special thanks to all stakeholders who participated in this study and to the Africa Tobacco Control Alliance (ATCA) and the African Center for Tobacco Industry Monitoring and Policy Research (ATIM) for their assistance with connecting the research team with their network of tobacco control stakeholders across Africa.

## Data availability statement

Data are available on reasonable request. Requests will be considered on a case-by-case basis.

## Declaration of Generative AI and AI assisted technologies in the writing process

The authors declare that no generative AI or AI assisted technology was used in the writing of this paper.

