## Supplementary materials SM1 and SM2 for "Exploring a roadmap to achieving tobacco endgame in sub-Saharan Africa: A qualitative study among stakeholders from 12 countries"

**Supplementary materials (SM)**

**SM1: Interview guide**

### Tobacco endgame strategies in Africa: Qualitative interview questions

Thank you for agreeing to participate in this study. I will be asking you a few questions to which I kindly request your honest opinion.

1. Please tell me about yourself and your work in tobacco control in your country
2. How would you describe the current state of tobacco use in Africa?
3. Are you familiar with the term “tobacco endgame”?
4. If yes, explain in your words what tobacco endgame is and its importance for tobacco control.
5. If no [interviewer explains what Tobacco endgame is all about], now that you know, what is the importance of tobacco endgame in tobacco control?
6. Are tobacco endgame strategies a matter of urgency in Africa and why?
7. Do you believe that unique African endgame strategies are required in order to end the use of tobacco in Africa, and why?
8. How important is it for Africa to have its own endgame strategies and why?
9. What strategies do you believe are currently being used to end tobacco use in Africa?
10. How successful do you think these strategies have been?
11. Would you say it is possible for Africa to end the use of tobacco in the continent? Give reasons for your answer.
12. What are the challenges faced by Africa in becoming a tobacco-free continent?
13. How can these challenges be overcome?
14. Besides the strategies that have been used, according to your knowledge, which other strategies can be used in Africa to end tobacco use?
15. Is the government doing enough to permanently eradicate nicotine/tobacco addiction in Africa?
16. What would you say is the role of the public in ending tobacco use?
17. What are the factors that will decide the success of tobacco endgame strategies in Africa?
18. What would you recommend to be done by African countries in order to reduce tobacco deaths and illnesses?

**SM2: Table S1**

**Number of participants invited and interviewed participants per country**

| **Country** | **Number of participants invited** | **Number of participants interviewed & included in the study** |
| --- | --- | --- |
| Algeria | 1 | - |
| Benin | 2 | 2 |
| Botswana | 1 | - |
| Burkina Faso | 6 | 2 |
| Burundi | 1 | - |
| Cameroon | 1 | - |
| Central African Republic | 1 | - |
| Chad | 2 | - |
| Democratic republic of Congo | 3 | - |
| Djibouti | 1 | - |
| Ethiopia | 3 | 2 |
| Gabon | 1 | - |
| Gambia | 1 | - |
| Ghana | 5 | 3 |
| Guinea | 1 | - |
| Ivory Coast | 1 | 1 |
| Kenya | 6 | 4 |
| Madagascar | 1 | - |
| Mali | 1 | - |
| Mozambique | 1 | - |
| Niger | 1 | - |
| Nigeria | 6 | 5 |
| Rwanda | 1 | - |
| Senegal | 4 | 2 |
| Sierra Leone | 2 | - |
| South Africa | 15 | 5* |
| Togo | 4 | 1 |
| Uganda | 2 | 1 |
| Zambia | 1 | - |
| Zimbabwe | 1 | 1 |
| **Total** | **77** | **29** |

*10 participants were interviewed but 5 declined speaking about regional concerns due to their limited knowledge about regional issues in tobacco control
